# Small Airway Disease as long-term Sequela of COVID-19: Use of Expiratory CT despite Improvement in Pulmonary Function test

**DOI:** 10.1101/2021.10.19.21265028

**Authors:** Alpana Garg, Prashant Nagpal, Sachin Goyal, Alejandro P. Comellas

## Abstract

**Background:** It is important to understand the spectrum of pulmonary diseases that patients are presenting after recovery from initial SARS-CoV-2 infection. We aim to study small airway disease and changes in Computed Tomography (CT)and pulmonary function tests (PFTs) with time.

**Methods:** This is retrospective observation study including adult patients with confirmed SARS-CoV-2 infection with at-least two CT scans either during acute (defined as <1 month) or subacute (1-3 months) or chronic (>3months) phase after positive test. Radiological features and follow up PFTs were obtained.

**Results:** 22 patients met the inclusion criteria with mean age 57.6 years (range 36-83). Out of these,18 (81.81%) were hospitalized. Mean duration of diagnosis to CT and PFT was 192.68(112-385) days and 161.54(31-259) days respectively. On PFTs, restrictive pulmonary physiology was predominant finding during subacute 56.25% (9/16) and chronic phases 47% (7/15). PFTs improved significantly with time {FEV1((p=0.0361), FVC (p=0.0341), FEF 25%-75% (p=0.0259) and DLCO (p=0.0019)}, but there was persistent air trapping in the expiratory chronic phase CT. There was resolution of ground glass opacity, consolidation, and bronchiectasis however air trapping increased with time in 41.61% (10/21) of subacute CTs compared to 81.25% (13/16) in chronic CTs.

**Conclusion:** Our study shows evidence of airway as well as parenchymal disease as relatively long-term sequel of SARS-CoV-2 infection. It also highlights the natural course and spontaneous recovery of some radiological and pulmonary function test abnormalities over time with evidence of persistent small airway disease (air trapping) on expiratory CT imaging months after infection.

**Highlights:**

1. Long term pulmonary complication of SARS-CoV-2 infection include small airway disease
2. There is role of inspiratory and expiratory Computed tomography (CT) scan to identify air trapping in patients with persistent respiratory symptoms after SARS-CoV-2 infection
3. Normal spirometry and normal routine CT may not be sufficient to characterize and identify cause of persistent respiratory complains in patients after COVID-19
4. This study highlights persistent parenchymal and physiological airway abnormalities more than six months after recovery from initial SARS-CoV-2 infection

## Introduction

Severe acute respiratory syndrome coronavirus 2 (SARS-CoV-2) has infected more than 196 million people and has resulted in more than 4 million deaths worldwide.^1^ Patients who have survived the acute illness are increasing and they are now presenting with persistent respiratory complains like dyspnea and cough, months after acute illness.^2^ Now known as Post-Acute Sequalae of SARS-CoV-2 (PASC) or long COVID, it is important to understand the spectrum of underlying respiratory diseases during this phase.^3^ Acute pulmonary complications of this respiratory virus include viral pneumonia, acute respiratory distress syndrome (ARDS), pulmonary embolism whereas long term complications include but not limited to persistent interstitial abnormalities, bronchiectasis and pulmonary fibrosis.^4-8^ Follow-up data from other studies report persistent physiological and parenchymal abnormalities, especially in hospitalized patients with severe initial SARS-CoV-2 infection.^9-11^ However, there are only limited number of studies reporting evidence of functional small airway disease (fSAD) after SARS-CoV-2 infection.^12,13^

Small airways are non-cartilaginous (less than 2 mm in diameter), start from generation 8th of lung branching and are often defined as ‘quiet zones’ of the lungs.^13,14^ Being small is diameter, they are susceptible to occlusion by small, inhaled toxins or pathogens (also known as post infectious bronchiolitis) or by local or systemic inflammatory damage with underlying conditions like connective tissue disease, post lung or bone marrow transplant and environmental or inhalational exposure (e.g. smoking).^13,15^ The diagnosis is difficult to establish as traditional lung function tests remain normal until late in disease course and direct assessment of small airways is beyond resolution of routine Computed Tomography (CT) scanners.^14^ However, small airways are indirectly evaluated on CT by subjective and quantitative measures of air trapping (or gas trapping) and mosaic attenuation especially in expiratory scans which suggests heterogenous distribution of airway disease.^13, 14, 16^

The aim of this study was to understand the natural course of radiological and pulmonary function tests, in a cohort of patients who presented to post-Covid-19 clinic and underwent longitudinal follow up with expiratory and inspiratory pulmonary imaging and pulmonary function test (PFT). The greater aim was to find the proportion of patients who had evidence of small airway disease based on imaging and pulmonary function tests.

## Materials and Methods

### Study Design

This study was a retrospective observational descriptive study conducted at University of Iowa hospitals & clinics. The study was approved by Institutional board review (IRB# 2020055421). The study included patients from inception of post covid clinic from June 1,2020 to July 15^th^,2021. Written informed consent was obtained from all the patients included in the study.

All adult patients who presented to outpatient post-COVID-19 clinic for follow up, had either a positive SARS-CoV-2 reverse transcriptase polymerase chain reaction (RT-PCR) or positive SARS-CoV-2 antibody test and had at least two temporal CT scans since the diagnosis and a PFT were included in the study. CT imaging was classified as acute CT (defined as CT < 1 month since COVID-19 diagnosis), subacute CT (1-3 months since COVID-19 diagnosis) or a chronic CT (> 3 months since COVID-19 diagnosis). The CT imaging protocol consisted of acquisition of lung imaging at inspiration (coached to total lung capacity) and expiration (coached to residual volume). The details of the CT imaging protocol used for the COVID-19 clinic patients is detailed elsewhere.^16^ All the patients who had prior documented history of lung fibrosis or interstitial lung disease (ILD) were excluded from the study.

### Data Collection

We collected demographics (age, sex, race, ethnicity, BMI, smoking history), past medical history, presenting complains, level of care received after the diagnosis [ambulatory or hospitalized or Intensive care unit (ICU)], length of stay, and treatment received after COVID-19 diagnosis. At presentation and follow-up in the post COVID-19 clinic we obtained inspiratory and expiratory CT and PFT. All the CT images were reviewed by experienced Cardiothoracic imaging fellowship trained radiologist. PFT done at clinic visit were reviewed by an experienced pulmonologist and further characterized as subacute (1-3 months after COVID-19 diagnosis) or chronic (>3 months after COVID-19 diagnosis). CT images were interpreted using standardized imaging nomenclature (such as ground-glass opacity [GGO], consolidation, air-trapping).^17^ All PFTs were obtained and interpreted using Standardized American Thoracic Society criteria.^18^ The PFTs were timed with either subacute or chronic CT scans. The following parameters were measured: forced vital capacity (FVC), forced expiratory capacity at first second of exhalation (FEV1), total lung capacity (TLC), residual volume (RV) and diffusion capacity of the lung for carbon monoxide (DLCO), Forced Expiratory Flow (FEF25%-75%). If hemoglobin values were available, then DLCO values were corrected for hemoglobin. PFT variables with values less than 80% were considered abnormal. Resting pulmonary hyperinflation was defined as RV/TLC ratio of more than or equal to 40%.

### Clinical Data Analysis

Primary outcomes were the clinical characteristics and abnormalities in the initial imaging and the follow up pulmonary imaging. Results were expressed using proportions, percentages, means and standard deviation or minimum and maximum values, as appropriate. Paired t-test was used to measure changes in FEV1, FVC and DLCO. A p-value less than 0.05 was considered statistically significant.

## Results

### Demographic and clinical characteristics

Out of twenty-two patients included in the study, most (18/22 i.e., 81.81%) were admitted to either medical floor (5/18) or the intensive care unit (13/18) and 4/22(18.18%) were managed outpatient during COVID-19 diagnosis. Majority of the patients were Caucasians (81.81%) and males with mean age of 57.6 years (range 36-83). Eleven out of 18 patients hospitalized, (61.11%) required oxygen at time of hospital discharge and 36.36% (8/22) underwent pulmonary rehabilitation. None of the patients were current smokers but 31.8% of patient smoked in the past with 26.42 mean pack years smoking history. The demographic and clinical characteristic of the patients is summarized in Table 1.

**Table 1.** Baseline characteristics of patients included in the study

| Characteristic | Total (n = 22) |
| --- | --- |
| Age, years(range) | 57.6 (36-83) |
| Gender |  |
| Male, n (%) | 18(81.81%) |
| Female, n (%) | 8(36.36%) |
| Race |  |
| Caucasian (%) | 18(81.81%) |
| African American, n (%) | 3(13.63%) |
| Native, n (%) | 1(4.54%) |
| Height, mean, in centimeter(range) | 172.69(157-190.5) |
| Weight, mean in Kilogram(range) | 98.00(62-147.9) |
| Body Mass Index, mean, Kg/m2(range) | 32.79(21.7-52.89) |
| Smoking |  |
| Current smoker | 0 |
| Past smoker, n (%) | 7(31.81%) |
| Past smoking (mean pack years) | 26.42 |
| Coexisting co-morbidities |  |
| Hypertension, n (%) | 14(63.63%) |
| Obesity, n (%) | 10(45.45%) |
| Diabetes Mellites, n (%) | 7(31.81%) |
| Asthma, n (%) | 3(13.63%) |
| Chronic Obstructive Pulmonary disease, n (%) | 3(13.63%) |
| Coronary artery disease, n (%) | 1(4.54%) |
| Congestive heart failure, n (%) | 3(13.63%) |
| Chronic kidney disease, n (%) | 2(9.09%) |
| Immunocompromised (organ-transplant, immunodeficiency), n (%) | 3(13.63%) |
| Connective tissue disease, n (%) | 1(4.54%) |
| Level of care received during acute COVID-19 |  |
| Inpatient at time of initial infection, n (%) | 18(81.81%) |
| Admitted to Medical ward, n (%) | 5 (22.72%) |
| Admitted to Intensive Care, n (%) | 13(59.09%) |
| Outpatient at time of initial infection, n (%) | 4(18.18%) |
| Mean Length of hospital stay, days | 27.5 |
| Treatment Received during acute COVID-19 |  |
| Supplemental oxygen through nasal cannula, n (%) | 2(9.09%) |
| Non-Invasive Mechanical ventilation, n (%) | 10(45.45%) |
| High Flow, n (%) | 9(40.9%) |
| Mechanical ventilation, n (%) | 7(31.81%) |
| Steroids, n (%) | 17(77.27%) |
| Remdesivir, n (%) | 9(40.9%) |
| Convalescent plasma, n (%) | 5(22.72%) |
| Antibiotics, n (%) | 13(59.09%) |
| Renal replacement therapy, n (%) | 3(13.63%) |
| Oxygen after hospital discharge [out of 18 patients hospitalized], n (%) | 11(61.11%) |
| Pulmonary rehabilitation, n (%) | 8(36.36%) |

### Pulmonary function tests and spirometry

The mean duration of follow up pulmonary function test from positive SARS-CoV-2 test was 161.54 (range 31-259) days. Spirometry was obtained during the subacute phase in 72.72% (16/22) patients. From these patients, 56.25% (9/16) had restrictive lung physiology and only 12.5% had obstructive lung physiology (2/16). In subacute phase, DLCO was measured in 50% (11/22) of the patients, out of which 72.72% (8/11) had reduced DLCO. Spirometry was obtained during the chronic phase in 68.18% (15/22) of patients, with 47% (7/15) having normal spirometry, while 33% (5/15) remained restrictive, and 20% (3/15) obstructive. DLCO was measured in 63.63% (14/22), with 50% of patients with reduced DLCO. This DLCO reduction suggests alveolar capillary defect. In addition, a group of 7 patients out of 22 patients had both spirometry measured during the subacute and chronic phases. As shown in Figure 1, FEV1((p=0.0361), FVC (p=0.0341) and FEF 25%-75% (p=0.0259) improved significantly with time. Four patients in this group had also DLCO measured during these time points, with significant change(p=0.0019). These results demonstrate improvement in lung function without any therapeutic intervention (including steroids).

**Figure 1.**
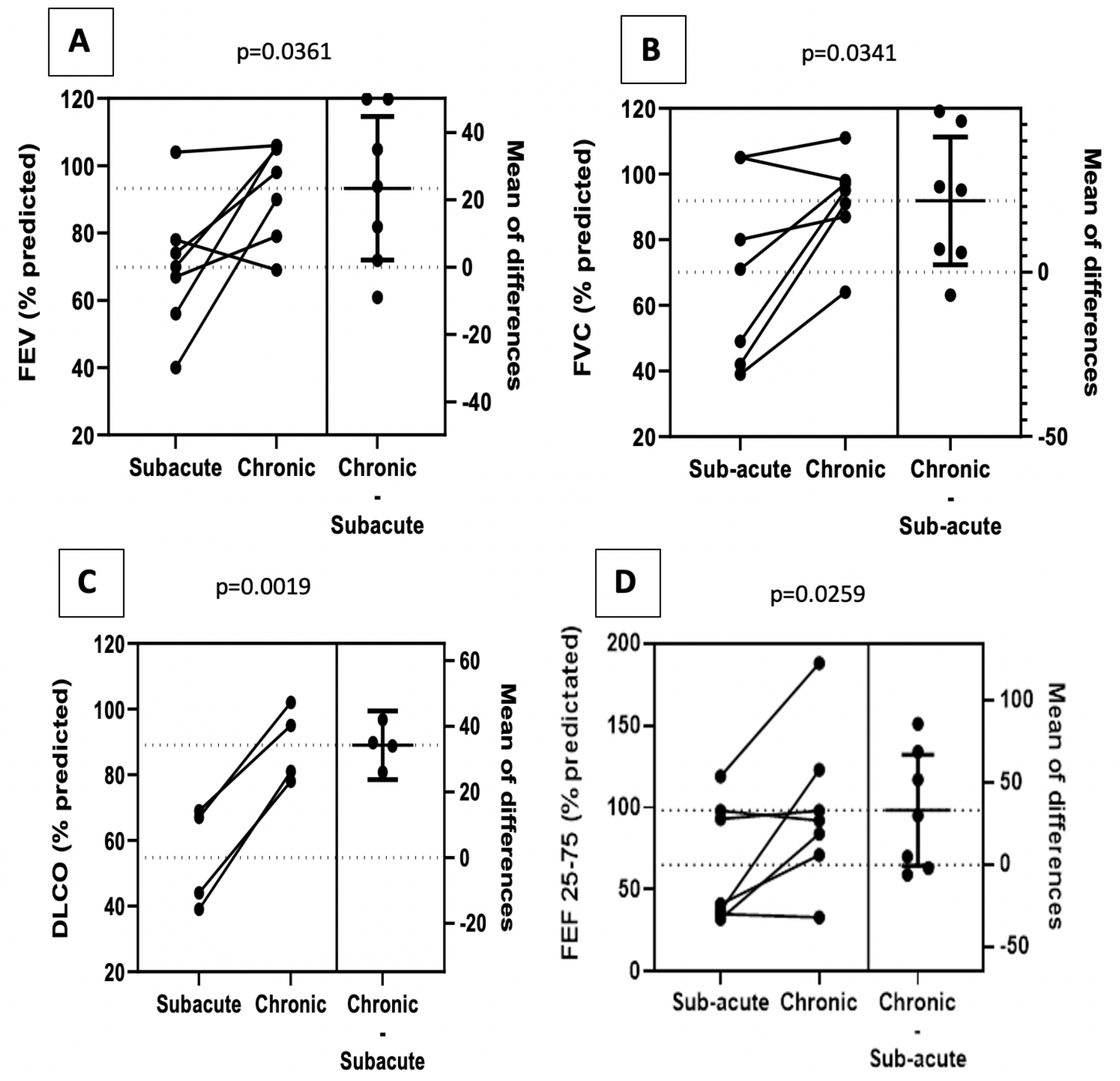
Improvement in FEV1 (A), FEV1 (B) and FEF 25-75% (D) in seven patients in subacute and chronic phase with T-test reaching statistical significance. C) Improvement in DLCO in 4 patients (out of 7 patients for which value was available

### Radiological characteristics

The quality of all CT images was evaluated by Radiologist and only images with excellent quality with no motion or good quality with mild motion were included in the study. The details regarding the CT findings are depicted with individual clinical characteristics in Table 2. Out of 17 CTs in the acute phase, 11 were CT angiogram (CTA), 2 were CT with contrast and 4 were CT without contrast. With risk of thromboembolism during acute illness, we expected CTs done with contrast to look for pulmonary embolism. The mean duration of follow up for subacute CT was 55.75 days (31 to 90 days post-COVID 19) whereas, chronic CT was 192.68 days (range112-385). Most common finding in majority of acute phase and subacute phase CT was GGO: either only GGO or GGO associated with reticular interstitial thickening. All the (17/17) CTs done in acute phase and 95.23% (20/21) of CTs done in subacute phase had ground glass opacities. In the chronic phase CT, there was resolution of ground glass with persistent only in 43.75% (7/16).

**Table 2.** Computed Tomography (CT) characteristics of Patients at Initial and follow up scans

| Imaging Characteristics, n (%) | Lung imaging at less than 1 month of positive SARS-CoV-2 test<br>**Acute phase** | Lung imaging at 1-3 months of positive SARS-CoV-2 test<br>**Subacute phase** | Lung imaging at more than 3 months of positive SARS-CoV-2 test<br>**Chronic phase** |
| --- | --- | --- | --- |
| Total CT scans | 17 | 21 | 16 |
| Mean duration of imaging from SARS-CoV-2 test, days (minimum- maximum) | 19.76(1-30) | 55.75(31-90) | 192.68(112-385) |
| Lymphadenopathy | 6(35.29%) | 4(19.04%) | 1(6%) |
| Pleural Effusion | 1(5.88%) | 4(19.04%) | 1(6%) |
| Emphysema | 3(17.64%) | 3(14.28%) | 3(19%) |
| Bronchial wall thickening | 2(11.7%) | 1(4.76%) | 1(6.25%) |
| Ground glass opacity | 17(100%) | 20(95.23%) | 7(43.75%) |
| Ground glass opacity reticular | 14(82.35%) | 17(80.9%) | 8(50%) |
| Consolidation | 12(70.5%) | 3(14.2%) | 0 |
| Architectural distortion or scarring or Honeycombing | 3(17.64%) | 10(47.61%) | 6(37.5%) |
| Bronchiectasis | 4(23.52%) | 13(61.90%) | 7(43.75%) |
| Inspiration Quality (Good or excellent) | 17(100%) | 20(95.23%) | 16(100%) |
| Expiration Quality (Good or excellent) | 7(41.16%) | 12(57.14%) | 14/16 |
| Air Trapping | 4(23.52%) | 10(47.61%) | 13(81.25%) |
| Disease severity based on CT |  |  |  |
| Mild | 2(11.76%) | 4(19.04%) | 4(25%) |
| Moderate | 4(23.52%) | 9(42.85%) | 1(6.25%) |
| Severe | 11(64.70%) | 6(28.57%) | 2(12.5%) |
| No disease | 0 | 2(9.52%) | 8(50%) |
| Nodules |  |  |  |
| -Nodules Present (%) | 6(35.29%) | 8(38.09%) | 4/16(25%) |
| -Type |  |  |  |
| Solid | 3/7 | 3/8 | 2/4 |
| Both solid and GGO | 3/7 | 3/8 | 2/4 |
| Ground glass | 0/7 | 2/7 | 0/4 |

In acute phase CT, other pertinent findings included nodules 35.29% (6/17), mediastinal lymphadenopathy 35.29% (6/17) and pulmonary embolism in 17.64% (3/17).

In subacute phase, the most common CT modality was without contrast with inspiratory and expiratory phase (17/21) and 4 were CTA. Majority of CT imaging in subacute and chronic phases were done as per inspiratory/expiratory COVID-19 protocol for patients in post COVID-19 clinic. In chronic phase, all the CTs were done without contrast with inspiratory and expiratory phases. There was evidence of air trapping in 81.25% (13/16) CTs in chronic phase. When compared to acute phase CT, air trapping was present in 23.52% (4/17) and subacute 47.61% (10/21). Besides air trapping, there was evidence of bronchiectasis in 61.90% (13/21) of subacute phase CTs and 43.75% (7/16) in chronic phase CT suggesting slight improvement with time. Interestingly, architectural distortion and some parenchymal scarring was present in 47.61% (10/21) of subacute CTs and 37.5% of CTs in chronic phase. The individual patient characteristics and corresponding CT findings in subacute and chronic CT are highlighted in Supplementary Table 3. Also figure 2, 3 and 4 show representative examples of air trapping evident in expiratory CT images.

**Figure 2.**
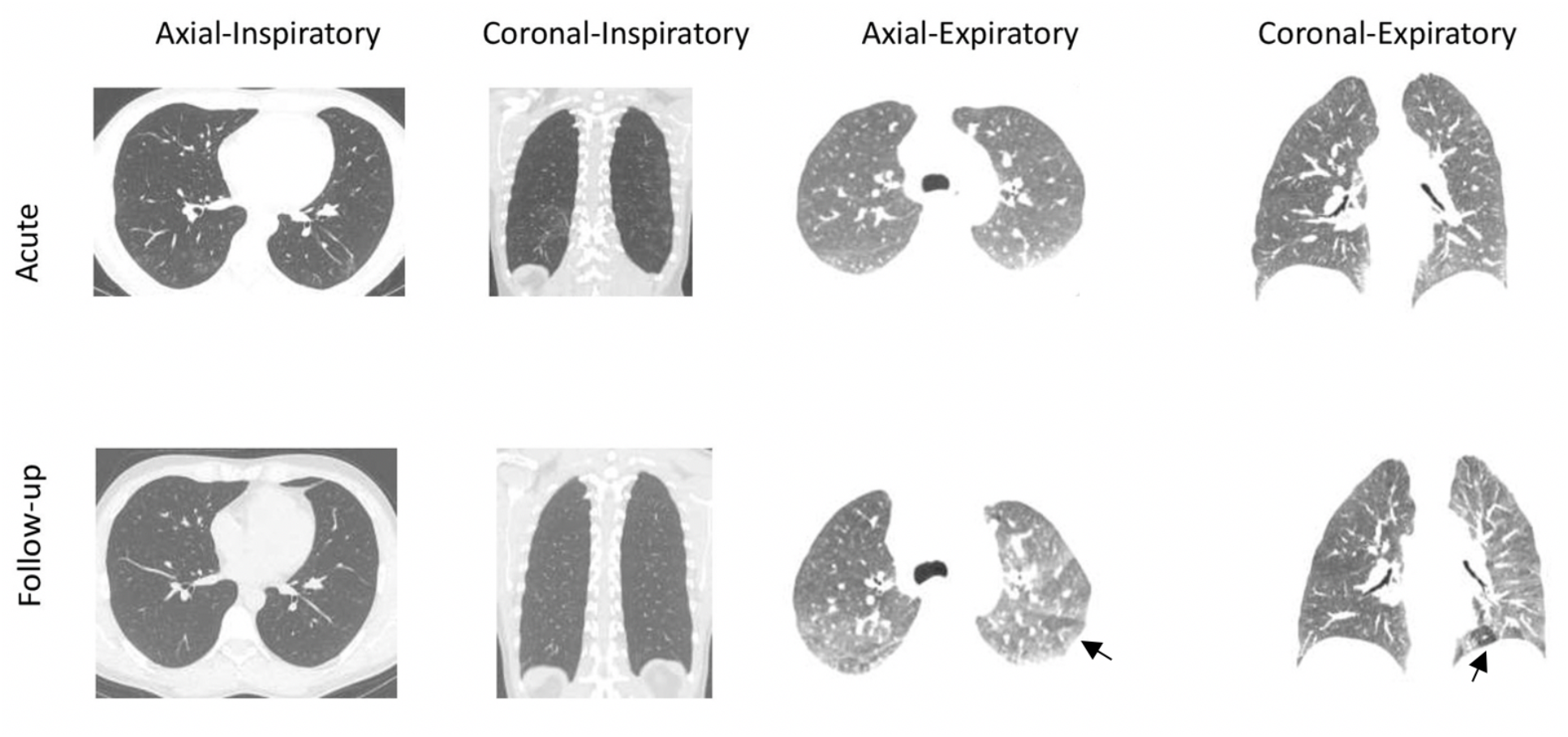
Middle aged man in age range 35-40 years (Case 5) previously healthy, non-smoker, mildly symptomatic with myalgias, chills and fever during initial infection with initial CT (top) at 30 days after positive SARS-CoV-2 test showing no significant air trapping. Follow up CT at 7 months showing mosaic attenuation and air trapping in expiratory films (arrows pointing towards air trapping)

**Figure 3.**
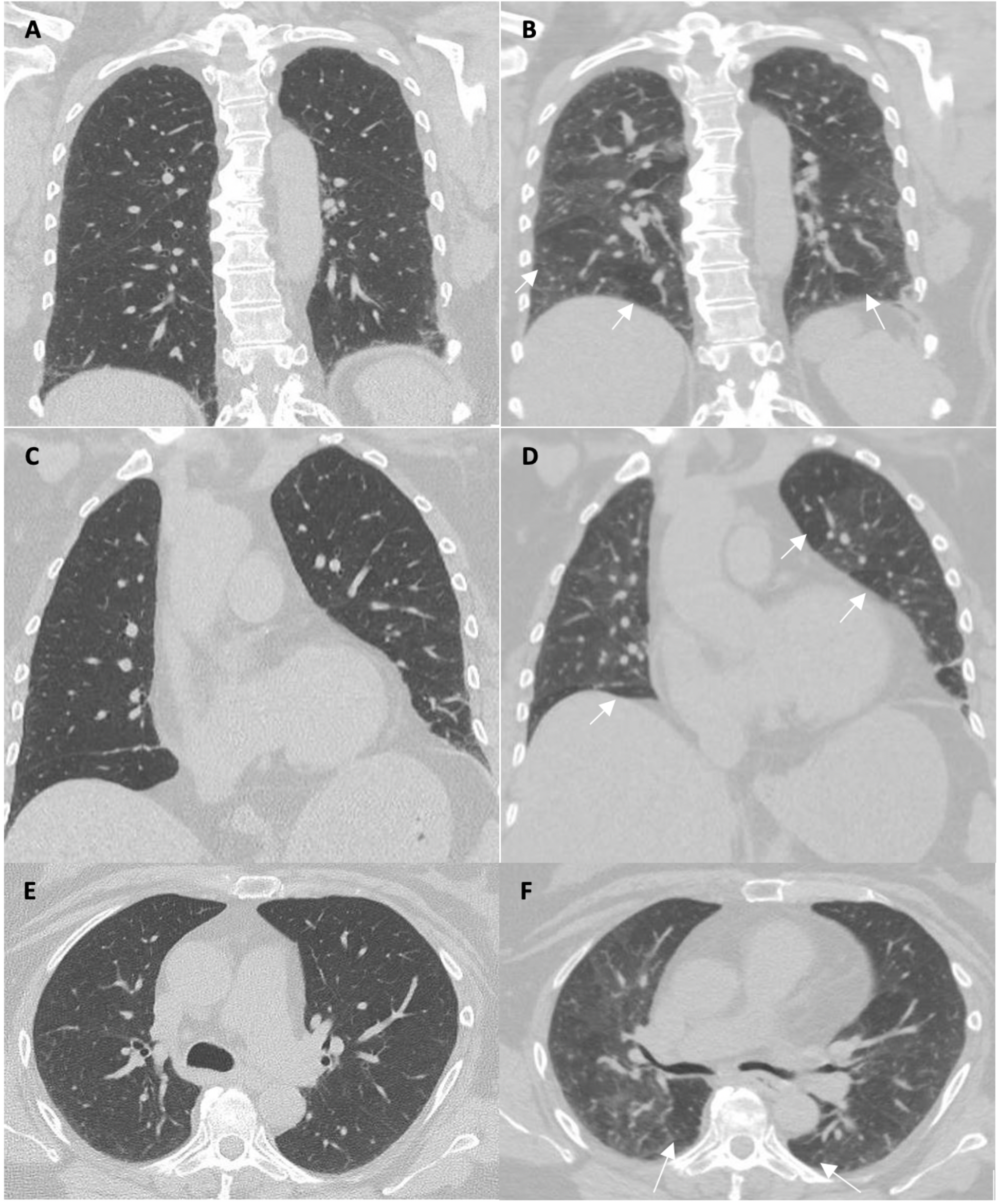
A woman in her 60s (Case 12) with history of hypertension and coronary artery disease admitted to medical floor with fever, cough, dyspnea for five days. She did not require oxygen during this time. She had persistent cough and dyspnea at follow up 230 days with CT inspiratory films (A, C, E) and expiratory films (B, C, D) showing bilateral air trapping at multiple areas (white arrows pointing to some areas) in expiratory films which was not present in the acute CT.

**Figure 4.**
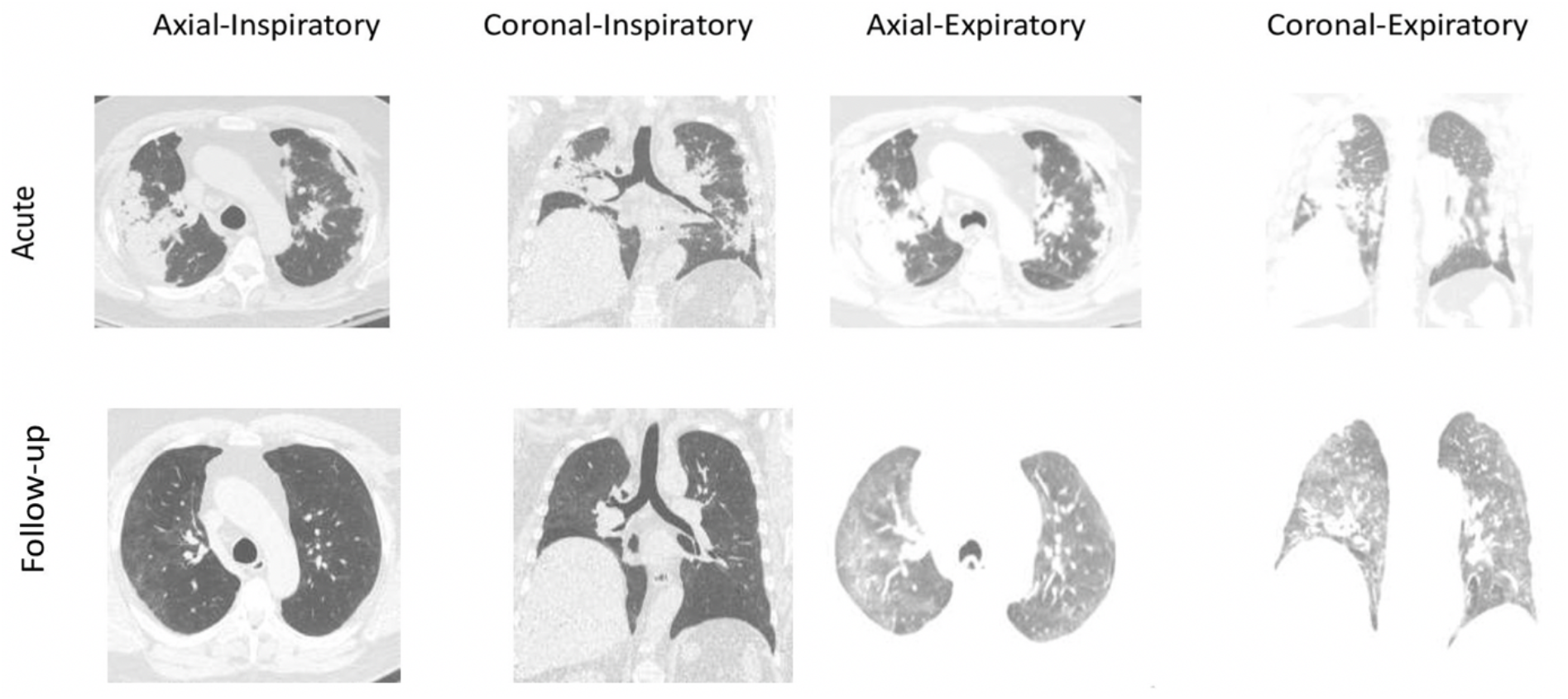
A man in his 60s (Case 8) presented with dyspnea admitted to critical care for respiratory failure requiring high flow oxygen due to severe COVID-19, hospitalized for 17 days, and discharged on oxygen. Acute CT (1 day after positive COVID-19) shows severe pneumonia with consolidation and ground glass opacities in all 5 lobes. Follow up CT at 207 days showing near complete resolution with bilateral air trapping (best seen in coronal section left lower lobe)

## Discussion

In this study we report that PFT changes including FEV1, FVC, FEF25-75%, DLCO, and lung parenchymal changes, such as GGO, bronchiectasis, architectural distortion, and even parenchymal scarring can improve over time. However, the proportion of patients with air trapping increased from the acute phase (23.52%) to the chronic phase (81.25%). The results of our study are in line with our previous study of 100 patients who presented for follow up after SARS-CoV-2 infection, with air trapping and evidence of small airway disease in 25.5% of patients in ambulatory group (not admitted to hospital), 34.5% in hospitalized group and 27.2% in intensive care group regardless of severity of infection.^13^ Recently, Huang et al reported air trapping in early stages of SARS-CoV-2 infection disease which persisted at two months follow up.^12^ This was a multicenter study done in China, included 108 patients, and found evidence of small airway disease in expiratory and inspiratory high-resolution CT.

The overall contribution of small airways is minimum to airway resistance and previous spirometry data suggests that spirometry abnormalities may not present until > 75% of airways are obstructed.^15, 19^ In our cohort, the spirometry changes and some parenchymal CT findings improved with time, but some parenchymal CT findings persist, and air trapping progressed over time. Without having acquired expiratory chest CT scans, we would have missed the high prevalence of air trapping despite improvement in pulmonary functions and lung parenchymal changes. Furthermore, none of the patients received long term systemic steroids, but there was spontaneous resolution of lung parenchymal changes including bronchiectasis and scarring with time, even in the patients with severe disease (Figure 4). The long-term clinical significance of these findings is yet unclear but raises the possibility that patients with COVID-19 infection can develop features of pulmonary obstructive lung disease due to damage to the small airways with potential to develop long-term bronchiolitis obliterans. As large number of patients with post-acute sequalae of SARS-CoV-2 (PASC) present with persistent respiratory complains, it will be important to understand the different phenotypes and tailor therapies so that unnecessary treatments with potential side-effects can be avoided. We would also emphasize that follow-up with CT imaging should be done carefully as subjecting patients to repeated radiological exposure and assessment of small airway is not feasible unless clinically indicated.^14^ While the objective evaluation of air trapping on CT is possible, qualitative assessment of air-trapping is highly subjective.^13, 16^ Moreover, single point interpretation of air-trapping is difficult as it is a non-specific sign and can be seen with some other physiological (e.g., obesity) and pathological conditions (e.g., pulmonary vascular congestion).^14, 20, 21^ However, temporal changes after COVID-19 as highlighted in this study can provide understanding of small airway disease change in terms of progression or improvement.

Our study highlights the natural course of the SARS-CO-V2 infection and effects on lungs. Our cohort was heterogeneous and included patients with variety of clinical characteristics and disease severity describing the evolution of image characteristics and spirometry with time. In this study, all the CT findings were evaluated by a single experienced fellowship trained radiologist, hence excluding inter-observer variability. The quality of expiratory and inspiratory films was assessed, and only studies with good or excellent quality CT were included. We followed the patients with inspiratory and expiratory CT scans and over more than six months follow-up (and one patient with 385 days follow up) and found the evidence of persistent (and even increasing) air trapping despite resolution (or stability) of other changes.

Our study had certain limitations. It is a single center, observational study which included relatively small number of symptomatic patients. However, since COVID-19 is a relatively new disease our aim was to include patients with comparable follow-up. Due to retrospective design, not all the patients were followed at the same regular intervals. Hence, there are some missing data points (not all the patients had imaging or PFT in all phases [acute, subacute, and chronic]). We cannot correlate the findings with underlying co-morbidities and clinical course as the sample size is small. Due to heterogeneous cohort, it will be difficult to determine which patients are more prone to developing small airway disease based on their clinical characteristics and illness course during acute infection. More longitudinal follow up studies including large number of patients can be done in future to delineate the risk factors and prevalence of fSAD in patients after SARS-CoV-2 infection. Also, studies are needed to understand the long-term consequences and treatment modalities in patients presenting with differing clinical and radiological phenotypes.

## Conclusion

Our study shows evidence of airway as well as parenchymal disease months after initial SARS-CoV-2 infection. It also highlights the natural course and spontaneous recovery of acute radiological and pulmonary functions abnormalities over time but indirect evidence of persistent (with temporal progression) small airway disease with air trapping months after COVID-19. Therefore, it is important to appropriately follow up patients after recovery from SARS-CoV-2 with respiratory complains by inspiratory and expiratory CT imaging even if pulmonary function tests are normal.

## Data Availability

All data produced in the present work are contained in the manuscript

## Conflicts of interest

None

## Acknowledgement

We would like to thank the patients who consented to participate in the study and supported research related to COVID-19

## Funding

None

## Author Contributions

AG was responsible for conceptualization, data curation, writing original draft

PN was responsible for reviewing the data, writing - review & editing.

SG was responsible for data collection, review and editing.

APC was responsible for conceptualization, formal analysis, methodology, supervision, writing - review & editing.

## Notes

### Competing Interest Statement

The authors have declared no competing interest.

### Funding Statement

This study did not receive any funding

### Author Declarations

IRB of University of Iowa gave ethical approval for this work

