## Supplemental Table 1 for "Small Airway Disease as long-term Sequela of COVID-19: Use of Expiratory CT despite Improvement in Pulmonary Function test"

Table 3 (Supplementary) Individual patient characteristics and summary of findings

| Case no. | Age Range | Sex  F=female  M=Male | A=Ambulatory/H=Hospitalized/ ICU=Intensive care unit | Relevant Clinical details | Days from positive SARS-CoV-2 test | Air trapping in subacute CT scan | Bronchiectasis or scarring or Architectural distortion in Subacute CT scan | Days from positive SARS-CoV-2 test | Air trapping in chronic CT | Bronchiectasis or scarring or Architectural distortion in Chronic CT | Days from PFTs | PFTs (OVD=obstructive ventilatory defect, RVD=restrictive ventilatory defect, Decreased DLCO, RV/TLC> 40%) |
| --- | --- | --- | --- | --- | --- | --- | --- | --- | --- | --- | --- | --- |
| 1. | 75-80 | F | ICU | Past smoker  Emphysema  persistent cough, SOB  Required MV | 90 | no | no | 288 | no | no | 116 | Obstructive ventilatory defect RV/TLC 45% |
| 2. | 65-70 | M | ICU | HTN, DM, asthma  Required high flow | 51 | No | yes | - | - | - | 359 | Normal |
| 3 | 35-40 | F | A | Previously healthy non-smoker  Persistent cough at follow-up | 52 | yes | yes | 385 | yes | yes | 283 | Normal |
| 4 | 45-50 | M | ICU | HTN, DM, post renal transplant  Required high flow  Cough, SOB, at follow up | 50 | yes | yes | 255 | yes | no | 239 | Restrictive ventilatory defect |
| *5 | 35-40 | M | A | Previously healthy, non -smoker  Cough, dyspnea, and increased air trapping at follow up | 31 | yes | no | 216 | no | yes | 34 | normal |
| 6 | 35-40 | F | ICU | DM  Cough and dyspnea at follow up | 61 | No | yes | - | - | - | 61 | RVD |
| 7 | 80-85 | F | H | - No previous history, had Pulmonary embolism acute phase | 37 | yes | yes | 219 | yes | yes | 22 | RVD, Decreased DLCO, RV/TLC 54% |
| *8 | 60-65 | M | ICU | - Obesity (BMI 41.8) was on high flow was on oxygen at discharge - Persistent cough and dyspnea at follow up | - | - | - | 207 | yes | yes | 32 | RVD, Decreased DLCO |
| 9 | 40-45 | M | ICU | - Required ECMO with tracheostomy later, oxygen at discharge persistent cough and dyspnea at follow up later evidence of fibrosis at follow up | 85 | yes | yes | - | - | - | 281 | RVD, Decreased DLCO |
| 10 | 60-65 | F | ICU | - Asthma, was on high flow, required oxygen at discharge, persistent cough, and dyspnea, GGO resolved, and bronchiectasis developed over time | 45 | yes | yes | 255 | yes | yes | 303 | At 4 months RVD, DLCO reduced  10 months-OVD, DLCO improved |
| 11 | 60-65 | F | A | - HTN, DM, cough, and dyspnea at follow up Atypical unilateral and non - peripheral GGO | 90 | yes | no | - | - | - | 85 | OVD, Early airway obstruction, RV/TLC 52% -->42% (Improved at 2 month), DLCO normal |
| *12 | 60-65 | F | H | - HTN, CAD, past smoker | 48 | - | no | 230 | yes | no | 230 | Normal |
| 13 | 65-70 | F | H | - Obesity ( BMI 53), COPD | 79 | yes | yes | - | - | - | 81 | RVD  DLCO reduced |
| 14 | 65-70 | M | ICU | - Severe COVID at baseline which improved on follow up | 35 | - | yes | 112 | yes | yes | 57 | RVD  Reduced DLCO |
| 15 | 70-75 | M | H | - HTN, CHF, ESRD had pulmonary embolism in acute phase - Required high flow and oxygen at discharge - Persistent dyspnea at follow-up | 88 | yes | yes | - | - | - | 165 | RVD  Reduced DLCO  RV/TLC 49% |
| 16 | 60-65 | M | ICU | HTN, DM, Obesity (BMI 40)  Required high flow and NIMV, oxygen at discharge, pulmonary rehabilitation  Persistent dyspnea, Scarring and severe disease with air trapping which became more prominent with time | 34 | no | yes | 128 | yes | yes | 226 | RVD  Reduced DLCO  RV/TLC 42% |
| 17 | 60-65 | M | H | HTN  Dyspnea at follow up | 54 | yes | no | 112 | yes | no | 216 | Normal, Reduced DLCO |
| 18 | 50-55 | M | ICU | HTN, ESRD, COPD, transplant COVID with emphysema with improvement over time | 62 | - | yes | 129 | yes | yes | 119 | RVD  Reduced DLCO |
| 19 | 45-50 | M | ICU | HTN  Required oxygen at discharge and pulmonary rehabilitation | 46 | - | yes | 145 | no | yes | 238 | RVD, Reduced DLCO |
| 20 | 55-60 | M | ICU | HTN  Required oxygen at discharge and pulmonary rehabilitation  Subpleural lines and bronchiectasis | 37 | - | yes | 113 | yes | yes | 113 | RVD  Reduced DLCO |
| 21 | 40-45 | M | ICU | DM, Obesity (BMI-49.77)  Required oxygen at discharge and pulmonary rehabilitation  complete clearing of subpleural disease | 34 | - | no | 166 | no | no | 49 | Normal |
| 22 | 60-65 | M | A | HTN, immune deficiency  Complete clearance of GGO but air trapping in lower lobes | 40 | - | no | 123 | yes | no | 125 | OVD, RV/TLC 49% |

Footnotes

1. Columns marked with hyphen (-) for which imaging was not available
2. (*) for cases 5, 8 and 12 with detailed images Figure 2,3 and 4

Abbreviations-PFTs- Pulmonary Function tests, forced vital capacity (FVC), forced expiratory capacity at first second of exhalation (FEV1), total lung capacity (TLC), residual volume (RV) and diffusion capacity of the lung for carbon monoxide (DLCO), Forced expiratory Flow (FEF_25%-75%_)
